# COVID-19 related outcomes for hospitalised older people at risk of frailty

**DOI:** 10.1101/2020.11.16.20232447

**Authors:** Rob Konstant-Hambling, Towhid Imam, Rhiannon K Owen, Andrew Street, Laia Maynou, Suzanne Arkill, Simon Conroy

## Abstract

**Background:** The COVID-19 pandemic has had a disproportionate impact upon older people. Frailty is being used to further refine the risk of poor outcomes in hospitalised older people. But studies to date on COVID related outcomes using frailty scales have reported inconsistent findings. We plan a retrospective cohort study using national data sources across England. The objectives are:

1. To determine if there is there an association between COVID-19 infection (virus identified), frailty risk (measured using the Hospital Frailty Risk Score - HFRS) and all-cause mortality.
2. To evaluate the association between HFRS in people with COVID-19 infection (virus identified), and the following secondary outcomes: hospital length of stay, critical care (entry to critical care, critical care length of stay and deaths in or following a critical care stay).
3. To determine if there is there an association between COVID-19 infection (virus identified), frailty risk (measured using the HFRS) and costs captured using Healthcare Resource Group tariffs.

**Methods:** This will be a retrospective cohort study using the NHS England Secondary Uses Service (SUS) electronic database, which records hospital activity and outcomes for all patients admitted to National Health Service hospitals in England. The analyses will use data relating to the index hospital presentation, this being the individual’s first emergency presentation during the study period for which they received a COVID-19 test. The primary and secondary outcomes will be constructed for the index admission. The analyses will control for differences in individual characteristics, using a set of risk adjusters including frailty, age, sex, ethnicity, deprivation, Charlson Comorbidity Index, number of previous admissions, number of (surgical) procedures, Ambulatory Care Sensitive Conditions (ACSCs) and COVID-19 status.

**Results:** Baseline characteristics will be reported using descriptive statistics. Mortality will be described using survival analysis, displayed as Kaplan Meier plots. A Cox proportional hazards model using robust standard errors to account for multiple observations (arising from readmissions) of the same individual will be fitted. The analyses will control for differences in individual characteristics, using a set of risk adjusters including frailty (Hospital Frailty Risk Score (HFRS)), age, sex, ethnicity, deprivation, Charlson Comorbidity Index, number of previous admissions, number of (surgical) procedures, Ambulatory Care Sensitive Conditions (ACSCs) and COVID-19 status (ICD-10 codes). Adjusted and unadjusted hazard ratios will be used to compare the rate of death for those with and without confirmed COVID-19, at different HFRS levels. We will test for an interaction between COVID-19 status and HFRS. A logit model will be implemented to analyse the secondary outcomes of admission to critical care mortality at 30 days, and mortality in critical care. For length of stay in hospital and in critical care, Poisson or negative binomial regression models will be fitted depending upon the dispersion.

**Impact:** The results of the study will inform clinicians about how best to use the frailty concept when assessing older people with COVID-19, for example in national guidelines that the study team have been involved in preparing: https://www.criticalcarenice.org.uk/.

## Background

The COVID-19 pandemic has challenged health and social care systems globally. A particular concern has been the availability of sufficient critical care capacity to support those individuals with severe respiratory complications following COVID-19 infection^1^. This has led to countries around the world thinking about how best to allocate potentially limited resources; an emerging feature of such considerations has been to use the frailty construct^2^. Frailty is a state of increased vulnerability to poor resolution of homoeostasis after a stressor event, which increases the risk of adverse outcomes, including delirium, disability and death^3 4 5^.

Where frailty has been studied in the critical care context, lower levels of frailty are associated with better outcomes^6-10^. However, most studies have used the frailty construct as a dichotomous measure rather than as a scale^11^, as per the original intended use – to recognise the heterogeneity of older people^12 13^. A common feature of the pandemic response globally has been to impose national and/or local lockdowns, and to shield populations perceived to be especially vulnerable to COVID-19. As a consequence, many countries have seen a profound reduction in presentations to urgent care settings, which may impact upon the discrimination of existing validated frailty scales.

Our recent systematic review [submitted] identified 16 studies assessing the influence of frailty on COVID-19 related mortality in hospitalised patients. The overall quality of the studies was reasonable, but the more robust studies showed that in older people hospitalised with COVID-19 infection that frailty (measured using the Clinical Frailty Scale) is a predictor of mortality. However, this was not consistent across all cohorts, with some showing a more complex interaction between frailty and COVID-19 status: two studies with contemporaneous non-COVID controls, found a sub-additive interaction with frailty i.e. that the mortality seen in severely frail older people was not as high as expected and excess mortality in those relatively fitter.

The aim of this study will be to examine COVID-19 outcomes across all of England, using the Hospital Frailty Risk Score (HFRS)^14^.

## Methods

The objectives of the study are:

- To determine if there is there an association between COVID-19 infection (virus identified), frailty risk (measured using the HFRS) and all-cause mortality (primary outcome).
- To evaluate the association between HFRS in people with COVID-19 infection (virus identified), and the following secondary outcomes: hospital length of stay and critical care use (entry to critical care, critical care length of stay and deaths in or following a critical care stay).
- To determine if there is there an association between COVID-19 infection (virus identified), frailty risk (measured using the HFRS) and costs captured using Healthcare Resource Group tariffs.

### Study design

This will be a retrospective cohort study using the NHS England Secondary Uses Service (SUS) electronic database.

### Data sources

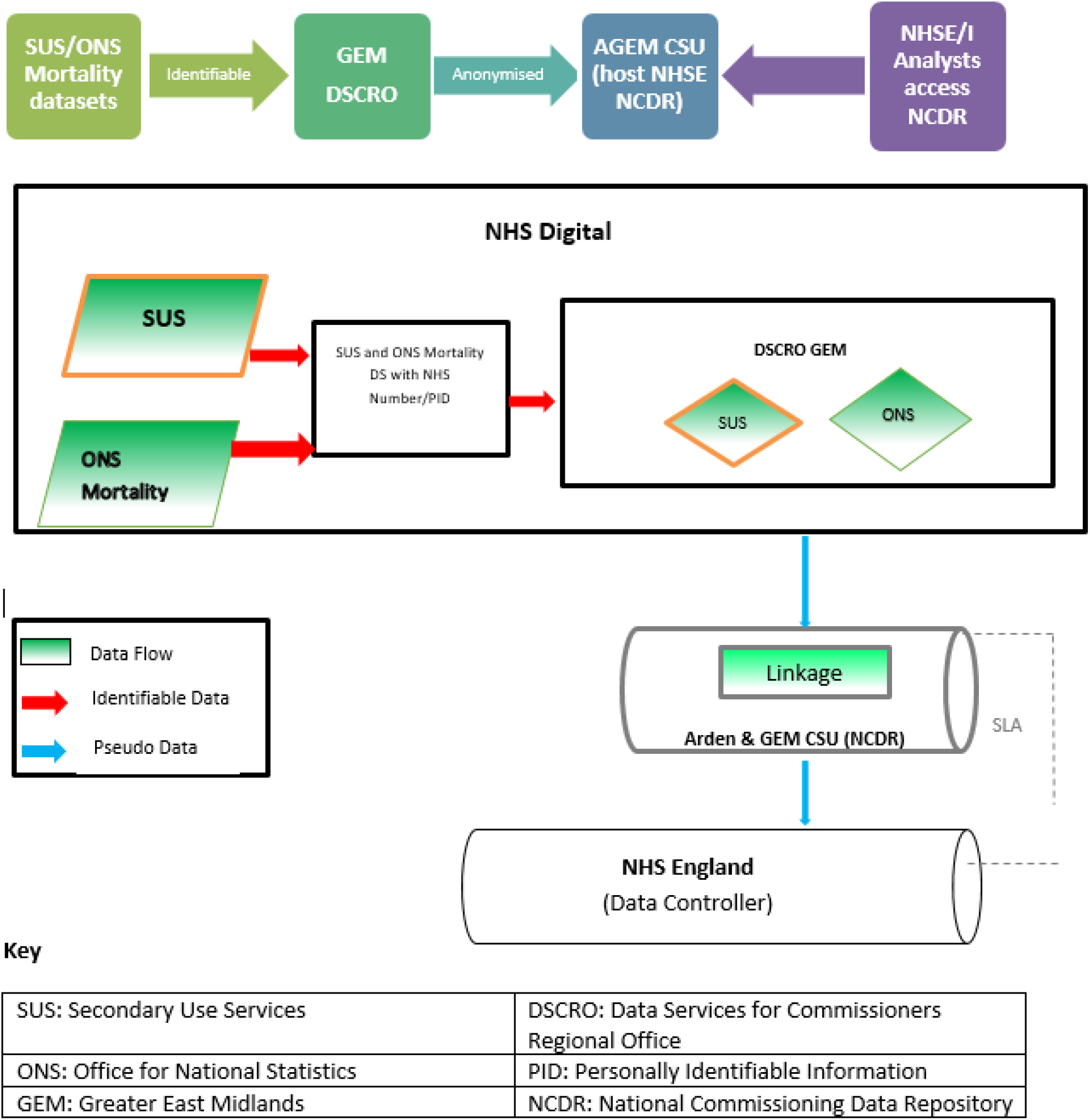

### Inclusion/exclusion criteria

Participants will be included if they are 75 years or older and have clinically suspected and test confirmed COVID-19 infection OR if they have clinically suspected but test negative COVID-19 infection, defined by ICD-10 codes (see below).

Participants will be excluded if they do not have essential information to undertake linkage.

### Analyses

#### Primary endpoint/outcome

The primary outcome will be all-cause mortality captured by the Office for National Statistics (ONS). ONS receive reports of all deaths recorded by death certificates, which have been validated by a medical examiner to check their accuracy, for all deaths in England. It includes deaths within as well as outside of hospital. We will report short term mortality (at 30 days) and mortality up until the study end date.

#### Secondary endpoints/outcomes

The secondary outcomes for this study will relate to proximal hospital outcomes including hospital length of stay, critical care use (entry to critical care, critical care length of stay and deaths in critical care) and mortality up until the study end-date (31^st^ August 2020).

Hospital length of stay will be calculated using ‘Finished Consultant Episodes) (FCE). Each FCE represent a period of time under the care of a hospital specialist; these can be summed together to provide the overall length of stay in hospital (in days), or specific spells under certain specialties, such as critical care.

Critical care use: SUS dataflows include a specific extension which captures critical care activity (from the Critical Care Minimum Dataset (CCMDS)); common identifiers allow this data to be linked to the admissions data described above at both patient and admission level.

Baseline characteristics will be reported using descriptive statistics. Mortality will be described using survival analysis, displayed as Kaplan Meier plots. Individuals alive at study end will be censored on 31^st^ August 2020. For individuals with readmissions, all preceding admissions will be censored the day before subsequent readmission. A Cox proportional hazards model using robust standard errors to account for multiple observations (arising from readmissions) of the same individual will be fitted. For subsequent admissions without further COVID-19 testing, a last-test carried forward approach will be implemented. The assumption of proportionality for Cox proportional hazards model will be assessed for COVID-19 status and HFRS categories. Adjusted and unadjusted hazard ratios will be used to compare the rate of death for those with and without confirmed COVID-19, at different HFRS levels. We will test for an interaction between COVID-19 status and HFRS. The magnitude of missing data will be thoroughly explored and where appropriate, multiple imputation will be considered. Statistical significance will be assessed at the 5% significance level.

A logit model will be implemented to analyse the secondary outcomes of admission to critical care mortality at 30 days, and mortality in critical care. For, length of stay in hospital and in critical care, Poisson or negative binomial regression models will be fitted depending upon the dispersion. Descriptive statistics will be calculated as unadjusted and adjusted odds or rates.

#### Risk adjustment data definitions

Of key focus in this study is the association between frailty and the primary and secondary outcomes for COVID-19 patients. Frailty will be measured using the HFRS, which was validated in people aged 75 or more who had been admitted to an acute hospital. The HFRS uses ICD-10 codes pertaining elective or non-elective hospital admissions to generate a frailty risk score. The HFRS uses diagnostic information in an algorithm that identifies the risk of frailty and outcomes such as death or unplanned hospital readmissions^14^. In the national validation cohort (n=1,013,590), compared with the 42% patients with the lowest risk scores, the 20% patients with the highest HFRSs had increased odds of 30-day mortality (odds ratio 1.71; 95% CI 1.68–1.75), long hospital stay (6.03; 5.92–6.10), and 30-day readmission (1.48; 1.46–1.50). The c-statistics between individuals for these three outcomes were 0.60, 0.68, and 0.56, respectively. The HFRS offers an opportunity to assess frailty as a case-mix characteristic; its relative ease of application makes it an ideal tool for use in national datasets to provide a population perspective. The HFRS will be applied to people aged 75 or older, admitted between February 2020 to July 2020. The SUS database allows searching of any previous admission (in this case, over the preceding two years) to identify any of the ICD-10 codes used to generate the HFRS. The original HFRS validation described three categories of low (<5), intermediate (5-15), and high frailty risk (>15), which will be applied for this study. Building upon ongoing work, we will draw on diagnostic information from the patient’s current hospital admission and data from the previous two emergency admissions that occurred within one year of the current admission to construct the HFRS.

#### Other risk adjustment variables

- Age will be captured in years and gender as male or female.
- Ethnicity is recorded in the SUS record according to one of 18 pre-specified categories.
- Deprivation is captured using the Index of Multiple Deprivation (IMD) score, derived from participants’ postcodes, reported as deciles.
- The Charlson Comorbidity Index (CCI) is a widely used measure of comorbidities, using ICD-10 codes for selected conditions to create a weighted score which is an important predictor of mortality in the acute hospital setting^15^. The CCI will be calculated for the incident admission only.
- The number of previous admissions is calculated as the number of completed secondary care episodes in the previous year prior to study entry.
- The number of procedures reflects surgical procedures undertaken during the index admission, as a case-mix adjuster.
- Ambulatory Care Sensitive Conditions (ACSC) defined as per the NHS digital method (https://digital.nhs.uk/data-and-information/data-tools-and-services/data-services/innovative-uses-of-data/demand-on-healthcare/ambulatory-care-sensitive-conditions#top).
- COVID-19 status will be defined by ICD10 codes U07.1 ‘COVID-19, virus identified’ and U07.2 ‘U07.2 COVID-19, virus not identified’ recorded in the SUS record. U07.1 is used when COVID-19 has been confirmed by laboratory testing and U07.2 when a clinical diagnosis of COVID-19 was suspected, but where laboratory confirmation is inconclusive or not available.

Our analyses will take account of the hospital in which patients are treated, given the possibility that outcomes may be partly related to differences in how hospitals organise care. This will be achieved by specifying fixed or random effects regression models that recognise the multi-level structure of the data, with patients clustered within hospitals^16-18^.

Acknowledging that those admitted at weekend are more likely to die than those admitted during the week^19^, we shall include day of admission among the explanatory variables.

Finally, our analysis will recognise that treatment of COVID19 has changed continually since patients were first admitted with the condition, and so we would expect those admitted more recently to have better outcomes. To capture this, our models will include a running time variable indicating how many weeks after the first week of February when the patient was admitted.

### Data handling and record keeping

No patient identifiable data will be held at the University of Leicester, nor will any individual patient data be seen by any of the University employees.

The study statisticians/econometricians (Owen, Maynou & Street) will supervise the NHS England analyst (RKH) through regular Microsoft Teams meetings, reviewing and guiding on the Stata codes that will be used to undertake the analysis and checking the results emerging on an iterative basis. They will at no stage be able to view any patient identifiable data or pseudonymised data; their contact will only be with RKH.

#### Data Handling

The organisation which will be receiving personal data is GEM DSCRO, which is part of NHS Digital. Under the Health and Social Care Act 2012 NHS Digital is designated as the national NHS Safe Haven for receiving personal identifiable data. In addition, under the HSC Act 2012, Greater and East Midlands DSCRO, has the responsibility to receive all data. NHS Digital collects data, as per the Directions (2015) issued to it by NHS England.

Data processed by GEM DSCRO is anonymised in accordance with the NHS Digital DSfC Anonymisation Requirements for Data used for Commissioning Purposes (in line with the ICO Anonymisation Code of Practice) before being securely transferred to Arden and GEM Commissioning Support Unit (CSU), (which is part of NHS England), where NHS England’s National Commissioning Data Repository (NCDR) is hosted.

Under strict access controls, NHS England and NHS Improvement’s approved analysts will use remote access arrangements to query the pseudonymised record level data which is held within the NCDR in order for them to analyse the data. The data can be accessed remotely from multiple locations in England using secure VPN or the N3 network, depending on where NHS Analysts are based. Access is secured via two personal user IDs and passwords; one to login in the terminal services server giving access to the Arden GEM network domain and then a further login into the SQL Server environment where the user is given read-only access to the data.

NHS England and NHS Improvement has implemented strict access controls to limit the amount of pseudonymised data which is made available to analysts. In order for users to access data on the NCDR they are required to outline in their access request application the purposes for which they require access.

Once access to data on the NCDR is granted, according to the role and user requirements, access is secured by using 2 factor authentification via VPN and on the N3 network. NHS England and NHS Improvement believe that the wider benefits of using the data to meet its statutory duties to ensure that patients receive the most appropriate care outweigh the extremely low risk of re-identification from the processing activities required.

This data will be linked to SUS data on a pseudonymised NHS Number. Where there is a request to link any two datasets the NCDR team, carry out a number of checks on the data requested to ensure that the risk of re-identification is minimised and, where applicable, there is also small number suppression. Where the risk of re-identification is deemed to cross the threshold of tolerance, the NCDR team carry out the data query of the datasets and produce an aggregated report.

Rob Konstant-Hambling is a NHS England analyst who works within the Arden & GEM CSU safe haven. RKH will undertake the analysis within the safe haven (according to the protocol prepared by the research team). Only summary results will be shared with the research team – no patient identifiable information will leave the safe haven at any stage, and RKH as the analyst will only ever see pseudonymised data, no patient identifiable data. No data will be store data the University. NHS data remains stored on NHS systems as per NHS standard operating procedures. **No** University employees will have access to the data for analysis, monitoring auditing or inspection.

For a fuller description of the data handling processes and the legal basis for accessing eth data, please see Appendix 1.

## Data Availability

If anyone wanted to replicate the analyses, they would have to request access from the data owner/controller (NHS England). The study protocol will be made available, as will the Stata codes for the analysis.

## Ethical and regulatory considerations

This study will be conducted in full conformity with the current revision of the Declaration of Helsinki (last amended October 2000, with additional footnotes added 2002 and 2004) and ICH Guidelines for Good Clinical Practice (CPMP/ICH/135/95) July 1996.

Fulham Research Ethics Committee (part of the UK Health Regulatory Authority) reviewed the application on and issued a Favourable Opinion (reference 289267). The University of Leicester acted as study sponsor (reference 0804 COVID & Frailty).

## Dissemination

The results of the study will inform clinicians about how best to use the frailty concept when assessing older people with COVID-19, for example in national guidelines that the study team have been involved in preparing: https://www.criticalcarenice.org.uk/.

